# Prevalence and risk factors of internet gaming disorder and problematic internet use before and during the COVID-19 pandemic: A large online survey of Japanese adults

**DOI:** 10.1101/2021.03.30.21254614

**Authors:** Taiki Oka, Toshitaka Hamamura, Yuka Miyake, Nao Kobayashi, Masaru Honjo, Mitsuo Kawato, Takatomi Kubo, Toshinori Chiba

## Abstract

Internet gaming disorder (IGD) and problematic internet use (PIU) are becoming increasingly detrimental in modern society, with serious consequences for daily functioning. IGD and PIU may be exacerbated by lifestyle changes imposed by the coronavirus 2019 (COVID-19) pandemic. This study investigated changes in IGD and PIU during the pandemic and risk factors for them. This study is a part of a larger online study on problematic smartphone use in Japan, originally planned in 2019, and expanded in August 2020 to include the impact of COVID-19. 51,246 adults completed an online survey during the pandemic (August 2020), in Japan. Of these, 3,938 had also completed the survey before the onset of the pandemic (December, 2019) and were used as the study population to determine how the pandemic has influenced IGD and PIU. IGD was assessed using the Internet Gaming Disorder Scale (IGDS). PIU was measured using the Compulsive Internet Use Scale (CIUS). The prevalence of probable IGD during COVID-19 was 4.1% [95%CI, 3.9% to 4.2%] overall (N=51,246), and 8.6% among younger people (age < 30), higher than reported before the pandemic (1 - 2.5%). Probable PIU was 7.8% [95%CI, 7.6% to 8.1%] overall, and 17.0% [95%CI, 15.9% to 18.2%] among younger people, also higher than reported before the pandemic (3.2 - 3.7%). Comparisons before and during the pandemic, revealed that probable IGD prevalence has increased 1.6 times, and probable PIU prevalence by 1.5 times (IGD: *t*_3937_ = 5.93, *p* < .001, PIU: *t*_3937_ = 6.95, *p* < .001). Youth (age < 30) and COVID-19 infection were strongly associated with IGD exacerbation (odds ratio, 2.10 [95%CI, 1.18 to 3.75] and 5.67 [95%CI, 1.33 to 24.16]). Internet gaming disorder and problematic internet use appear to be aggravated by the pandemic. In particular, younger persons and people infected with COVID-19 are at higher risk for Internet Gaming Disorder. Prevention of these problems is needed.

## Introduction

The coronavirus 2019 (COVID-19) pandemic has affected all aspects of society (Holmes et al., 2020; McGinty et al., 2020). Previous studies have suggested that stressors due to the pandemic contribute to increased addictive behaviors, such as substance use, alcohol, food, and social media (Bonny-Noach and Gold, 2020; Panno et al., 2020). The World Health Organization (WHO) has warned that during the COVID-19 pandemic, screen time and game-playing time may increase. This increases the risk of Internet and gaming addiction(WHO, 2020). Increased video gameplay and internet use among young people have been reported(Schmidt et al., 2020), possibly because of pandemic-induced lifestyle changes (e.g., staying at home, quarantines, closed workplaces, and schools (King et al., 2020; WHO, 2020)). While video games and the internet provide entertainment and convenience, maladaptive engagement in these activities can lead to various mental health problems, including internet gaming disorder (IGD) and problematic internet use (PIU). IGD and PIU are associated with greater psychological distress, poorer sleep quality, and severe social withdrawal, known as *‘hikikomori’* (Kato et al., 2020; Wong et al., 2020; Fazeli et al., 2020). Several opinion papers have cautioned that increased severity of IGD and IPU during a pandemic could persist after its subsidence, which may prolong poor quality of life for those affected and may impose a heavy economic burden on society (King et al., 2020; Ko and Yen, 2020). Young people have an especially great risk of internet-related problems and psychological distress during the pandemic (Chen et al., 2021, 2020; Fazeli et al., 2020; Sun et al., 2020). Such epidemiological studies are important in formulating policies for prevention and early intervention. However, to the best of our knowledge, most previous studies have been employed a cross-sectional approach. One study, which included only adolescents, found increased rates of internet-related problem behaviors in March 2020, compared with November 2019 (Chen et al., 2021).

Therefore, changing IGD and PIU levels among adults before and during the pandemic remain unclear. Using online survey data from 51,246 adults, we investigated the prevalence of IGD and PIU. The influence of the COVID-19 pandemic on IGD and PIU was assessed by comparing online survey data from a subsample of 3,938 adults collected before and during the pandemic.

## Material and methods

### Participants and Procedures

This investigation was a part of a larger study on the association between problematic smartphone use and multidimensional psychiatric states that was initiated in 2019 and later expanded to examine the impact of COVID-19 (Figure 1). Details can be found in our previous work(Chiba et al., 2020). It was approved by the Ethics Committee of the Advanced Telecommunications Research Institute International (Japan).

**Figure 1.**
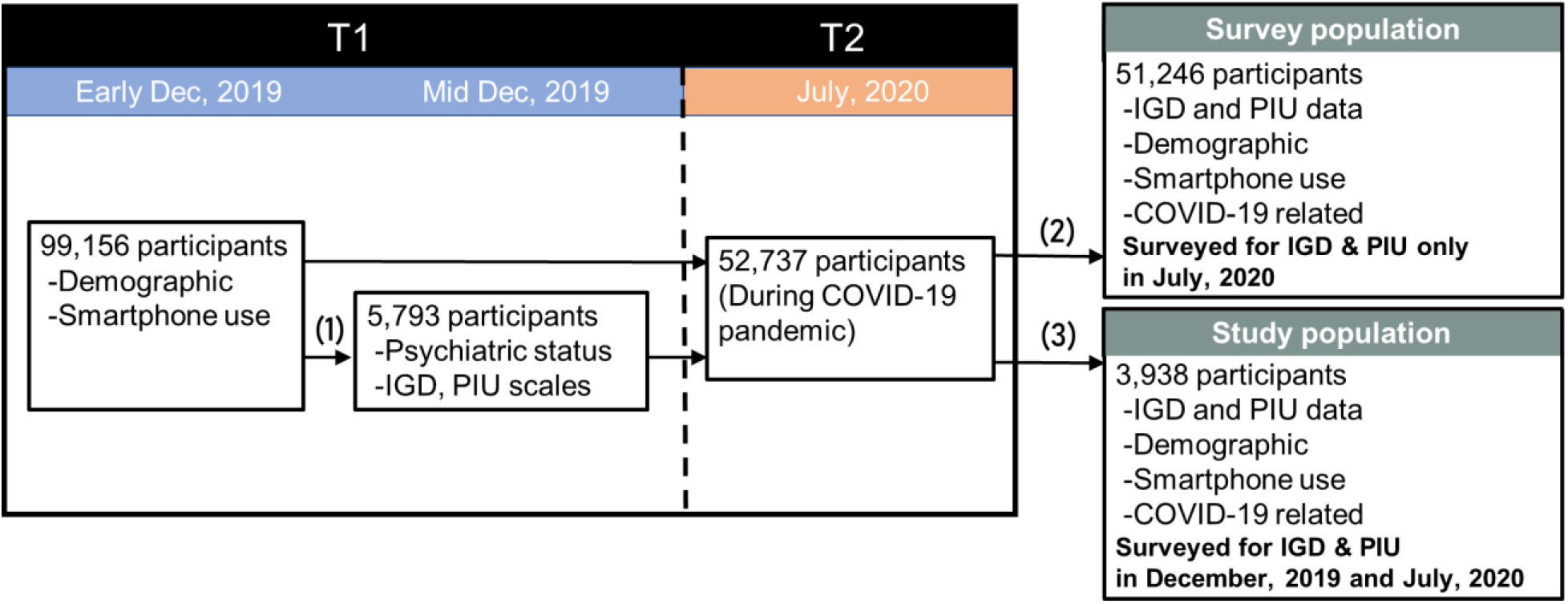
Flow chart showing numbers of participants and criteria for exclusion from the analyses. (1) This subpopulation includes equal numbers of individuals in each quintile relative to their problematic smartphone use score. These 5,793 respondents were drawn from the larger sample of 99,156 participants. (2) 52,737 participants from the early December, 2019 survey participated in a July, 2020 survey that included IGD/PIU and COVID-19 questions that were not in the original December, 2019 survey. 1,491 of the 52,737 participants were excluded because of inconsistencies in their answers. (e.g. sleep time = wake-up time, 1st gender ≠ 2nd gender). (3) 5,793 respondents from late December, 2019, who were asked questions about IGD/PIU, participated in the July, 2020 follow-up survey, in which they were also asked questions about COVID-19. 1,855 participants had to be excluded because they dropped out of the survey, there were inconsistencies in their answers, or because they responded identically to all items, using only the maximal or minimal values. This left 3,938 people who participated in both the late December, 2019 and July, 2020 surveys.

5,955 participants, screened in December 2019, immediately before COVID-19, reported their multi-dimensional psychiatric states, including IGD and CIUS (Figure 1). In order to assess changes in multiple psychiatric states during the pandemic, we conducted a follow-up survey in July 2020 that contained additional questions related to COVID-19 (e.g., Do you think that you have ever been infected by COVID-19?). 52,737 people responded to the July, 2020 survey, including 5,793 who participated in December, 2019 (Figure 1). Of the latter, 1,445 participants dropped out and 198 participants were excluded because of inconsistencies in their answers. An additional 212 participants were deleted because they responded identically to all items using only the maximum or minimum values. In the end, 3,938 respondents were included in the current analyses (Figure 1).

### Measures

IGD was measured according to the Japanese-version Internet Gaming Disorder Scale (IGDS), which consists of questions corresponding to each of the nine IGD symptoms defined in the Diagnostic and Statistical Manual of Mental Disorders, 5th ed (DSM-5)(American Psychiatric Association, 2013). Using a binary response format, items assessed the severity of each IGD symptom during the preceding 12 months. At least five symptoms are required to return a diagnosis of IGD. The reliability and validity of IGDS have been demonstrated with a Cronbach’s alpha of 0.93 (Lemmens et al., 2015). PIU was measured using the Compulsive Internet Use Scale (CIUS), which has a Cronbach’s alpha of 0.89. Construct validity has been confirmed by the strong positive correlation with the Online Cognition Scale (*r* = 0.70, *p* < .001) and the amount of time spent online (*r* = 0.33, *p* < .001) (Meerkerk et al., 2009). We defined probable IGD as a total IGDS ≥ 5, and probable PIU as a total CIUS score of ≥ 29 (Jeromin et al., 2016).

### Statistical analysis

Average IGDS and CIUS scores and the prevalence of probable IGD and PIU based on these scales were calculated for each age and sex group for both T1 (December, 2019, before COVID-19) and T2 (July, 2020, during COVID-19) (N=3,938). Changes in prevalence of probable IGD and probable PIU were analyzed using paired t-tests. We also analyzed each IGD item similarly. Bonferroni correction was used to adjust the multiple comparisons for analysis of each IGD item. Further, to explore the effects of demographic characteristics and infections of COVID-19 on IGD and PIU status, we conducted multiple logistic regression analyses to predict who developed probable IGD and probable PIU during the period between T1 and T2. To do so, participants without probable IGD or PIU at T1 were excluded for each analysis, separately.

Demographic characteristics included sex, marital status, the existence of children, age groups, household income, COVID-19 infection status, changes in the amount of communication with family from T1 to T2 (face-to-face and online, separately), and changes in smartphone use time from T1 to T2 (weekdays and weekends, separately).

#### Model specification

*logit (IGD or PIU) = Intercept + sex + marital status + the existence of children + age groups + household income + the status of COVID-19 infection + changes in the amount of face-to-face communication with family from T1 to T2 + changes in the amount of online communication with family from T1 to T2 + change in smartphone use time from T1 to T2 on weekdays + change in smartphone use time from T1 to T2 on weekends*

\**IGD and PIU* denote the state of probable IGD or probable PIU at T2, respectively

All demographic characteristics were treated as categorical variables. The odds ratio of each group for a given variable was calculated against the reference group. Reference groups were decided according to previous studies (Shen et al., 2020; Tsumura et al., 2018). To test potential selection bias, Pearson correlation analysis was performed for the prevalence of probable IGD and probable PIU in each age and sex group between the survey population and the study population. Statistical analyses were performed using Matlab version R2019b.

Statistical tests assumed a significance level α of 5%, except for the analysis of the difference of IGD items from T1 to T2, for which Bonferroni correction was used.

## Results

### Sample characteristics

The average age of participants was 46.6 years [standard deviation (SD) = 11.8], and 49.9% were male (Table 1). The prevalence of probable IGD was 4.1% (95%CI [3.9% to 4.2%]) and that of PIU was 7.8% (95%CI [7.6% to 8.0%]). The probable prevalence of IGD among young people (<30 age) was 8.6% (95%CI [7.8% to 9.5%]) and that of PIU was 17.0% (95%CI [15.9% to 18.2%]). Among adults, prevalence of probable IGD increased 1.5 times during the pandemic (*t*_3937_ = 5.93, *p* < .001) and PIU increased 1.6 times (*t*_3937_ = 6.95, *p* < .001). In young people, IGD prevalence increased 1.8 times during the pandemic (*t*_310_ = 3.36, *p* < .001) and PIU prevalence increased 1.6 times (*t*_310_ = 3.30, *p* = .001).

**Table 1.** Demographic characteristics in the survey population of the COVID-19 online survey.

|  |  | Sample size | Unweighted profile | Weighted profile | Mean IGDS score (95% CI) | Proportion with probable IGD (95% CI) | Mean CIUS score (95% CI) | Proportion with probable PIU (95% CI) |
| --- | --- | --- | --- | --- | --- | --- | --- | --- |
| **Total sample** |  | 51246 | 100% | 100% | 0.62 (0.61-0.63) | 4.1% (3.9-4.2) | 14.1 (14.1-14.1) | 7.8% (7.6-8.0) |
| **Age** | **Gender** |  |  |  |  |  |  |  |
| <20 | Male | 59 | 0.1% | 0.2% | 1.54 (0.99-2.10) | 13.6% (4.6-22.6) | 19.9 (17.4-22.4) | 15.3% (5.8-24.7) |
|  | Female | 172 | 0.3% | 0.2% | 0.97 (0.73-1.20) | 6.4% (2.7-10.1) | 21.3 (19.7-22.9) | 25.0% (18.5-31.5) |
| 20-29 | Male | 818 | 1.6% | 4.0% | 1.85 (1.69-2.01) | 16.5% (14.0-19.1) | 19.9 (19.2-20.7) | 19.4% (16.7-22.2) |
|  | Female | 3,129 | 6.1% | 3.7% | 1.04 (0.97-1.10) | 6.6% (5.7-7.5) | 19.1 (18.8-19.5) | 16.0% (14.7-17.3) |
| 30-39 | Male | 2,762 | 5.4% | 9.4% | 1.37 (1.29-1.44) | 10.3% (9.2-11.5) | 17.5 (17.1-17.9) | 13.9% (12.6-15.2) |
|  | Female | 6,657 | 13.0% | 9.0% | 0.75 (0.72-0.79) | 4.3% (3.8-4.8) | 17.6 (17.4-17.9) | 12.3% (11.5-13.1) |
| 40-49 | Male | 7,211 | 14.1% | 14.5% | 0.80 (0.76-0.84) | 6.2% (5.7-6.8) | 14.8 (14.6-15.0) | 8.8% (8.1-9.4) |
|  | Female | 7,472 | 14.6% | 14.2% | 0.53 (0.50-0.56) | 2.9% (2.5-3.2) | 14.7 (14.5-14.9) | 7.5% (6.9-8.1) |
| 50-59 | Male | 8,957 | 17.5% | 14.3% | 0.41 (0.39-0.44) | 3.0% (2.6-3.3) | 12.1 (11.9-12.3) | 5.2% (4.7-5.6) |
|  | Female | 5,662 | 11.0% | 14.2% | 0.39 (0.37-0.42) | 1.9% (1.5-2.2) | 12.2 (12.0-12.5) | 4.2% (3.7-4.7) |
| 60-69 | Male | 5,519 | 10.8% | 7.6% | 0.26 (0.23-0.28) | 1.4% (1.1-1.8) | 10.0 (9.8-10.3) | 2.3% (1.9-2.7) |
|  | Female | 2,470 | 4.8% | 8.0% | 0.26 (0.22-0.29) | 1.3% (0.8-1.7) | 9.9 (9.6-10.2) | 2.7% (2.0-3.3) |
| 70> | Male | 254 | 0.5% | 0.3% | 0.24 (0.12-0.36) | 2.0% (0.2-3.7) | 9.2 (8.2-10.2) | 3.1% (1.0-5.3) |
|  | Female | 104 | 0.2% | 0.4% | 0.38 (0.14-0.61) | 1.9% (-0.8-4.6) | 9.0 (7.4-10.6) | 1.9% (0-4.6) |

|  | Sample size | Unweighted profile | Mean IGDS score (95% CI) | Proportion with probable IGD (95% CI) | Mean CIUS score (95% CI) | Proportion with probable PIU (95% CI) |
| --- | --- | --- | --- | --- | --- | --- |
| **Employment status** | 51246 |  |  |  |  |  |
| Employee | 25041 | 49% | 0.67 (0.65-0.69) | 4.8% (4.5-5.0) | 14.4 (14.2-14.5) | 8.0% (7.6-8.3) |
| Executive | 1095 | 2% | 0.45 (0.38-0.53) | 2.7% (1.8-3.7) | 11.1 (10.6-11.7) | 3.9% (2.8-5.1) |
| Self-employment/Freelance | 3594 | 7% | 0.53 (0.49-0.58) | 3.8% (3.2-4.4) | 12.5 (12.2-12.8) | 5.9% (5.2-6.7) |
| Homemaker | 7731 | 15% | 0.54 (0.51-0.57) | 3.0% (2.6-3.4) | 14.4 (14.2-14.6) | 7.8% (7.2-8.4) |
| Part-time job | 8244 | 16% | 0.58 (0.55-0.61) | 3.3% (2.9-3.7) | 14.1 (13.9-14.3) | 7.0% (6.5-7.6) |
| Student | 833 | 2% | 1.26 (1.13-1.38) | 8.0% (6.2-9.9) | 21.2 (20.5-21.9) | 23.5% (20.6-26.4) |
| Other | 1510 | 3% | 0.62 (0.55-0.70) | 4.0% (3.0-5.0) | 14.1 (13.6-14.6) | 9.1% (7.7-10.6) |
| No employee | 3198 | 6% | 0.50 (0.46-0.54) | 2.8% (2.3-3.4) | 12.8 (12.5-13.2) | 7.5% (6.6-8.5) |
| **Existence of child(ren)** |  |  |  |  |  |  |
| No children | 21508 | 42% | 0.71 (0.69-0.73) | 4.5% (4.2-4.8) | 15.4 (15.2-15.5) | 9.9% (9.5-10.3) |
| Have child(ren) | 29738 |  | 0.55 (0.54-0.57) | 3.8% (3.5-4.0) | 13.2 (13.1-13.4) | 6.3% (6.1-6.6) |
| **Marital status** |  |  |  |  |  |  |
| Not married | 17664 | 34% | 0.72 (0.69-0.74) | 4.5% (4.2-4.8) | 15.5 (15.4-15.7) | 10.3% (9.9-10.8) |
| Married | 33582 | 66% | 0.57 (0.55-0.58) | 3.8% (3.6-4.0) | 13.4 (13.3-13.5) | 6.5% (6.2-6.8) |
| **Household income, \$** | | | | | | |
| ~39999 | 11962 | 23% | 0.65 (0.62-0.68) | 4.1% (3.8-4.5) | 14.3 (14.1-14.5) | 8.5% (8.0-9.0) |
| 40000~80000 | 17872 | 35% | 0.61 (0.59-0.63) | 4.0% (3.8-4.3) | 14.2 (14.0-14.3) | 7.7% (7.3-8.1) |
| 80000~120000 | 4687 | 9% | 0.62 (0.58-0.67) | 4.9% (4.3-5.5) | 13.6 (13.3-13.9) | 7.0% (6.3-7.8) |
| ~120000 | 4508 | 9% | 0.63 (0.58-0.68) | 4.7% (4.1-5.3) | 13.2 (12.9-13.5) | 6.8% (6.1-7.5) |
| Missing | 12217 | 24% | 0.59 (0.57-0.62) | 3.5% (3.1-3.8) | 14.4 (14.2-14.6) | 8.1% (7.6-8.6) |
| **Living with someone or alone** |  |  |  |  |  |  |
| Living with someone | 43249 | 84% | 0.61 (0.60-0.62) | 4.0% (3.8-4.2) | 14.1 (14.0-14.2) | 7.5% (7.3-7.8) |
| Alone | 7997 | 16% | 0.66 (0.62-0.69) | 4.3% (3.9-4.8) | 14.6 (14.3-14.8) | 9.4% (8.7-10.0) |
| **About COVID-19 infected** |  |  |  |  |  |  |
| Thinking negative themselves | 44360 | 87% | 0.56 (0.54-0.57) | 3.4% (3.2-3.5) | 14.1 (14.0-14.2) | 7.6% (7.3-7.8) |
| Received a diagnosis | 270 | 1% | 3.38 (3.03-3.73) | 39.3% (33.4-45.1) | 20.6 (19.4-21.8) | 19.6% (14.9-24.4) |
| Thinking positive themselves | 1136 | 2% | 1.41 (1.28-1.54) | 12.5% (10.6-14.4) | 18.7 (18.0-19.3) | 15.6% (13.5-17.7) |
| Missing | 5480 | 11% | 0.81 (0.77-0.86) | 6.1% (5.5-6.8) | 13.41(13.1-13.7) | 7.8% (7.1-8.5) |

Sample sizes are unweighted. Concerning gender and age subgroups, we used the weighted profile as representative of the Japanese population.
IGDS: Internet gaming disorder scale, CIUS: Compulsive internet use scale.
IGDS scores $\geq 5$ indicate a clinically probable level of IGD. CIUS scores $\geq 29$ indicate a clinically probable level of IGD.

### Changes of each IGD item between before and during the pandemic

Seven out of nine IGD symptoms increased significantly during the pandemic (Figure 2). Especially, ‘Tolerance’, ‘Withdrawal’, ‘Persistence’ and ‘Displacement’ were greatly exacerbated during the pandemic (Tolerance: *t*_3937_ *=* 4.60, *p* < .001; Withdrawal: *t*_3937_ *=* 4.44, *p* < .001; Persistence: *t*_3937_ *=* 5.40, *p* < .001; Displacement: *t*_3937_ *=* 5.94, *p* < .001).

**Figure 2.**
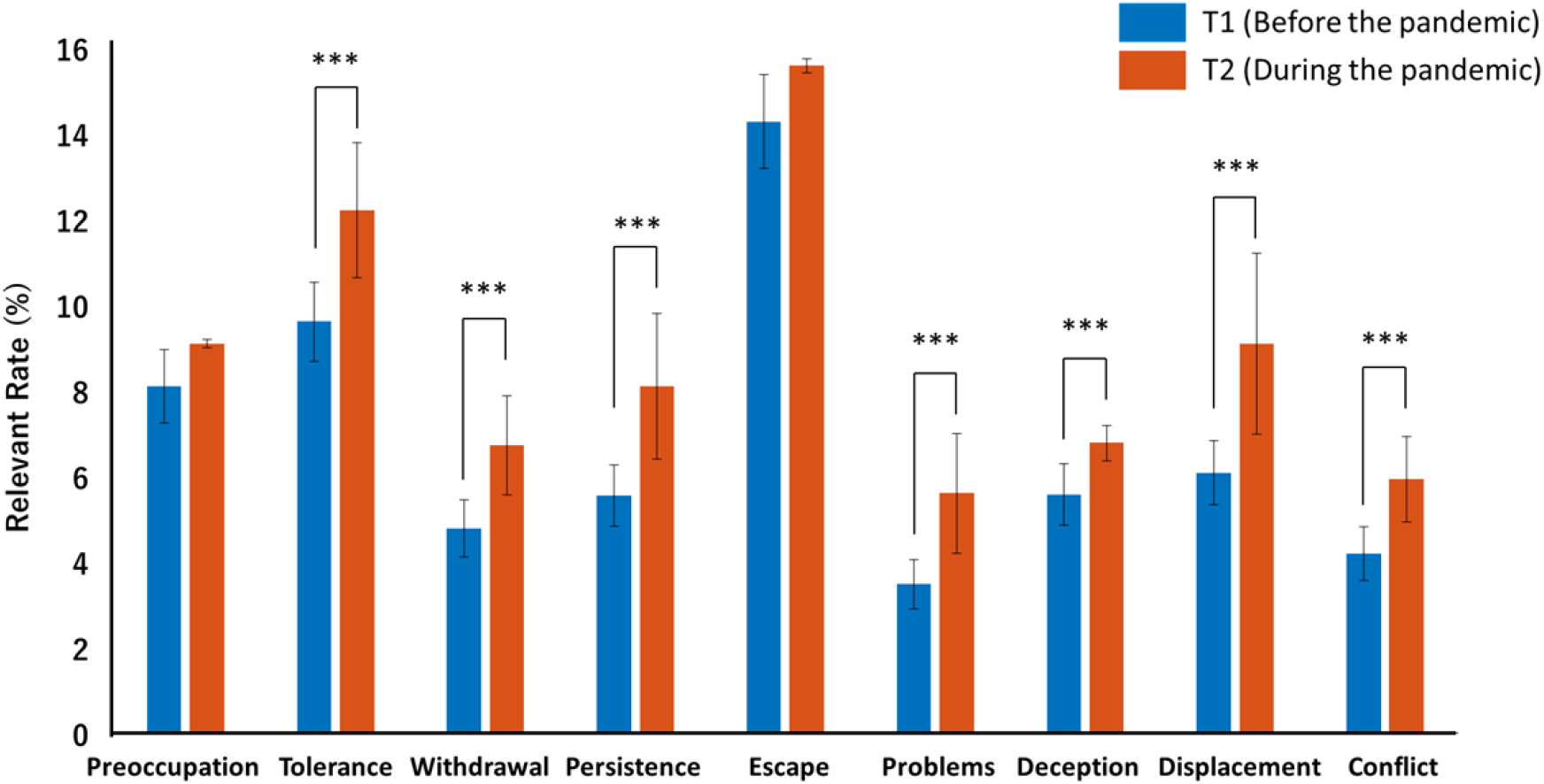
Comparison of IGD symptoms before and during the pandemic in the study population. These 9 symptoms are defined in DSM-5(American Psychiatric Association, 2013). *** *p≤* 0.001

### Multiple logistic regression analyses

Multiple logistic regression analyses were used to examine the effects of demographic factors on development of probable IGD and PIU (Table 2). The odds ratio (OR) for probable IGD development in males was 2.21 (95%CI [1.42 to 3.42]), whereas the OR among young people (<30) relative to people 40 to 49 years was 2.10 (95%CI [1.18 to 3.75]). The OR in those who decreased face-to-face communication-time with family was 1.71 (95%CI [1.15 to 2.53]) relative to those who reported no change in communication time. Also, the OR among those who decreased online communication time with family was 1.52 (95%CI [1.04 to 2.24]). The OR of those suffering COVID-19 infections was 5.67 (95%CI [1.33 to 24.16]) relative to those without COVID-19 infections.

**Table 2.**
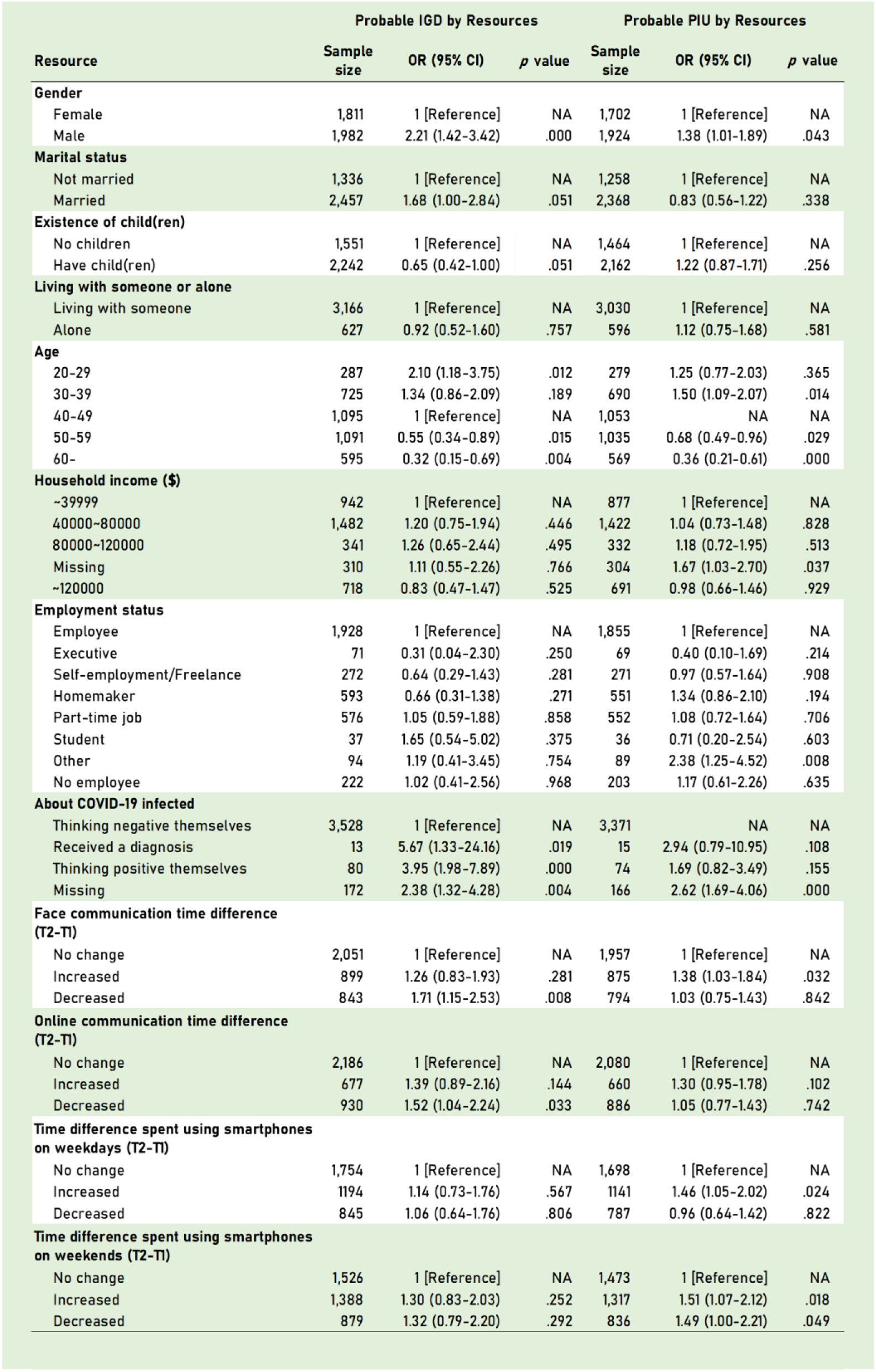
Results of logistic regression analysis predicting exacerbation of probable IGD and PIU (probable IGD development, N=3793; probable PIU development, N=3626).

| Resource | Probable IGD by Resources |  |  | Probable PIU by Resources |  |  |
| --- | --- | --- | --- | --- | --- | --- |
|  | Sample size | OR (95% CI) | p value | Sample size | OR (95% CI) | p value |
| **Gender** |  |  |  |  |  |  |
| Female | 1,811 | 1 [Reference] | NA | 1,702 | 1 [Reference] | NA |
| Male | 1,982 | 2.21 (1.42-3.42) | .000 | 1,924 | 1.38 (1.01-1.89) | .043 |
| **Marital status** |  |  |  |  |  |  |
| Not married | 1,336 | 1 [Reference] | NA | 1,258 | 1 [Reference] | NA |
| Married | 2,457 | 1.68 (1.00-2.84) | .051 | 2,368 | 0.83 (0.56-1.22) | .338 |
| **Existence of child(ren)** |  |  |  |  |  |  |
| No children | 1,551 | 1 [Reference] | NA | 1,464 | 1 [Reference] | NA |
| Have child(ren) | 2,242 | 0.65 (0.42-1.00) | .051 | 2,162 | 1.22 (0.87-1.71) | .256 |
| **Living with someone or alone** |  |  |  |  |  |  |
| Living with someone | 3,166 | 1 [Reference] | NA | 3,030 | 1 [Reference] | NA |
| Alone | 627 | 0.92 (0.52-1.60) | .757 | 596 | 1.12 (0.75-1.68) | .581 |
| **Age** |  |  |  |  |  |  |
| 20-29 | 287 | 2.10 (1.18-3.75) | .012 | 279 | 1.25 (0.77-2.03) | .365 |
| 30-39 | 725 | 1.34 (0.86-2.09) | .189 | 690 | 1.50 (1.09-2.07) | .014 |
| 40-49 | 1,095 | 1 [Reference] | NA | 1,053 | NA | NA |
| 50-59 | 1,091 | 0.55 (0.34-0.89) | .015 | 1,035 | 0.68 (0.49-0.96) | .029 |
| 60- | 595 | 0.32 (0.15-0.69) | .004 | 569 | 0.36 (0.21-0.61) | .000 |
| **Household income (\$)** | | | | | | |
| <39999 | 942 | 1 [Reference] | NA | 877 | 1 [Reference] | NA |
| 40000-80000 | 1,482 | 1.20 (0.75-1.94) | .446 | 1,422 | 1.04 (0.73-1.48) | .828 |
| 80000-120000 | 341 | 1.26 (0.65-2.44) | .495 | 332 | 1.18 (0.72-1.95) | .513 |
| Missing | 310 | 1.11 (0.55-2.26) | .766 | 304 | 1.67 (1.03-2.70) | .037 |
| >120000 | 718 | 0.83 (0.47-1.47) | .525 | 691 | 0.98 (0.66-1.46) | .929 |
| **Employment status** |  |  |  |  |  |  |
| Employee | 1,928 | 1 [Reference] | NA | 1,855 | 1 [Reference] | NA |
| Executive | 71 | 0.31 (0.04-2.30) | .250 | 69 | 0.40 (0.10-1.69) | .214 |
| Self-employment/Freelance | 272 | 0.64 (0.29-1.43) | .281 | 271 | 0.97 (0.57-1.64) | .908 |
| Homemaker | 593 | 0.66 (0.31-1.38) | .271 | 551 | 1.34 (0.86-2.10) | .194 |
| Part-time job | 576 | 1.05 (0.59-1.88) | .858 | 552 | 1.08 (0.72-1.64) | .706 |
| Student | 37 | 1.65 (0.54-5.02) | .375 | 36 | 0.71 (0.20-2.54) | .603 |
| Other | 94 | 1.19 (0.41-3.45) | .754 | 89 | 2.38 (1.25-4.52) | .008 |
| No employee | 222 | 1.02 (0.41-2.56) | .968 | 203 | 1.17 (0.61-2.26) | .635 |
| **About COVID-19 infected** |  |  |  |  |  |  |
| Thinking negative themselves | 3,528 | 1 [Reference] | NA | 3,371 | NA | NA |
| Received a diagnosis | 13 | 5.67 (1.33-24.16) | .019 | 15 | 2.94 (0.79-10.95) | .108 |
| Thinking positive themselves | 80 | 3.95 (1.98-7.89) | .000 | 74 | 1.69 (0.82-3.49) | .155 |
| Missing | 172 | 2.38 (1.32-4.28) | .004 | 166 | 2.62 (1.69-4.06) | .000 |
| **Face communication time difference (T2-T1)** |  |  |  |  |  |  |
| No change | 2,051 | 1 [Reference] | NA | 1,957 | 1 [Reference] | NA |
| Increased | 899 | 1.26 (0.83-1.93) | .281 | 875 | 1.38 (1.03-1.84) | .032 |
| Decreased | 843 | 1.71 (1.15-2.53) | .008 | 794 | 1.03 (0.75-1.43) | .842 |
| **Online communication time difference (T2-T1)** |  |  |  |  |  |  |
| No change | 2,186 | 1 [Reference] | NA | 2,080 | 1 [Reference] | NA |
| Increased | 677 | 1.39 (0.89-2.16) | .144 | 660 | 1.30 (0.95-1.78) | .102 |
| Decreased | 930 | 1.52 (1.04-2.24) | .033 | 886 | 1.05 (0.77-1.43) | .742 |
| **Time difference spent using smartphones on weekdays (T2-T1)** |  |  |  |  |  |  |
| No change | 1,754 | 1 [Reference] | NA | 1,698 | 1 [Reference] | NA |
| Increased | 1194 | 1.14 (0.73-1.76) | .567 | 1141 | 1.46 (1.05-2.02) | .024 |
| Decreased | 845 | 1.06 (0.64-1.76) | .806 | 787 | 0.96 (0.64-1.42) | .822 |
| **Time difference spent using smartphones on weekends (T2-T1)** |  |  |  |  |  |  |
| No change | 1,526 | 1 [Reference] | NA | 1,473 | 1 [Reference] | NA |
| Increased | 1,388 | 1.30 (0.83-2.03) | .252 | 1,317 | 1.51 (1.07-2.12) | .018 |
| Decreased | 879 | 1.32 (0.79-2.20) | .292 | 836 | 1.49 (1.00-2.21) | .049 |

The OR of people 30-39 years for probable PIU development relative to those aged between 40 and 49 was 1.50 (95%CI [1.09 to 2.07]). That of people who increased face-to-face-communication time with their families relative to those who reported no change was 1.38 (95%CI [1.03 to 1.84]). In regard to smartphone use, the OR of people who reported increases in their weekday smartphone use relative to that of those who reported no change was 1.46 (95%CI [1.05 to 2.02]). The OR of people who reported increased weekend smartphone use was 1.51 (95%CI [1.07 to 2.12]) while that of people who reported a decrease was 1.49 (95%CI [1.00 to 2.21]).

## Discussion

This is the first online survey to assess changes in prevalence of probable Internet gaming disorder (IGD) and problematic internet use (PIU) during the COVID-19 pandemic in Japan. Probable IGD prevalence was 4.1% during the pandemic in the survey population (N = 51,246). Analyses revealed that probable IGD prevalence increased more than 1.6 times (from 3.7% to 5.9%) during the pandemic. Probable IGD prevalence during the pandemic was higher than reported (1 - 2.5% (Pontes et al., 2016; Przybylski et al., 2017; Wu et al., 2018)) before the pandemic. Probable PIU prevalence was 7.8% during the pandemic in the survey population (N = 51,246) (Figure 1). Analyses of the study population (N = 3,938) revealed that probable PIU prevalence increased more than 1.5 times during (from 7.9% to 11.6%) the pandemic. Probable PIU prevalence was also higher than reported in studies before the pandemic (3.2 - 3.7% (Kuss et al., 2013a, 2013b)). Prevalence of probable IGD among younger people (<30 years old) was 8.6% in the survey population, higher than reported in studies among children and adolescents (1.2 - 7.5% (Jeong et al., 2020; Rehbein et al., 2015; Taechoyotin et al., 2020; Wartberg et al., 2020)) before the pandemic. It increased more than 1.8 times (from 7.7% to 13.8%) during the pandemic.

All symptoms of IGD increased during the pandemic. ‘Tolerance’, ‘Withdrawal’ and ‘Displacement’ were especially exacerbated. Rehbein (Rehbein et al., 2015) reported that symptoms related to ‘Tolerance’, ‘Withdrawal’, and ‘Displacement’ are the core symptoms of IGD. Moreover, these symptoms interfere with abstinence from gaming behavior (King et al., 2018), which prolongs the IGD state. Thus, the greater number of individuals with IGD is likely not just a transitory problem during the COVID-19 pandemic.

We also found that younger people (< 30) were at 2.1 times greater risk of IGD than the more mature group (40-49 years). This suggests that young people are more vulnerable to Internet-gaming-related problems. Decreased face-to-face and online communication time with family members were also associated with a higher risk of exacerbated IGD during the pandemic relative to persons whose screen time did not change (1.71 times and 1.52 times, respectively). These may suggest that loneliness or boredom due to decreasing time with family drove people to play games and become addicted. Additionally, we found that individuals infected with COVID-19 were at 5.67 times greater risk of progression in IGD than uninfected individuals. This suggests that stress and lifestyle changes caused by COVID-19 infection exacerbate IGD remarkably. Infected individuals reported an increase in IGD, possibly because they used the internet or games to avoid or cope with stress associated with the infection (Fazeli et al., 2020; Lee et al., 2017).

Increased smartphone use both on weekdays and weekends was associated with a greater risk of PIU development (weekdays: 1.46 times, weekends 1.51 times). Data from the Japanese Ministry of Internal Affairs and Communications shows increases in smartphone use on weekdays and weekends under states of emergency (“Information and Communications in Japan., 2020”). A previous study showed increases in smartphone use due to the pandemic (Chen et al., 2021). Our results may reflect forgoing non-essential and non-urgent activities due to the pandemic. On the other hand, those who decreased their smartphone use time on weekends still showed 1.49 times more PIU. This may reflect a shift from smartphone use away from home to use of personal computers or tablet computers at home. Therefore, simply monitoring smartphone use is not sufficient for early detection of PIU. Instead, we need to monitor screen time regardless of the device. People who increased face-to-face communication time with family were still at 1.38 times higher risk of PIU development relative to people with no change. This result is inconsistent with a previous study that reported face-to-face communication may protect against Internet-related behavior problems (Kim et al., 2015). Further studies are required to further illuminate the underlying basis for this inconsistency, such as differences in culture or momentum of the pandemic.

As shown above, most of the risks are common to both IGD and PIU, but some factors seem to have different effects on those risks. For example, a decrease in face-to-face communication time with family was a significant risk factor of IGD. In contrast, an increase in face-to-face communication time with family was a significant risk factor for PIU. Thus, approaches for early detection and prevention of internet-addictive behavior may differ in each group. This should be examined in future research.

There are several limitations to this study. First, this is a survey using an online recruiting method, and there may be some sampling bias. The study population (N=3,938) was extracted from the survey population (N=51,246) such that the study population includes equal numbers of individuals in each quintile relative to the problematic smartphone use score. However, Pearson correlations of the prevalence of probable IGD and probable PIU in each age and sex group between the survey population and the study population were significant (*r* = 0.94, *p* < .001, *r* = 0.90, *p* < .001). This shows the reliability of probable IGD and probable PIU as ratio scales, even if the raw value was overestimated. Second, this survey was taken in Japan.

Instead of locking down the city, the Japanese government declared a state of emergency to control the spread of the COVID-19 pandemic. People were encouraged, but not forced to stay home. Therefore, it is unclear whether the results of this Japanese study apply equally to other countries, especially those that locked down to control the pandemic. It is important to compare these results with data from other countries having different ethnicities and government strategies. Despite these limitations, this study shows increasing IGD and PIU and identifies at-risk populations for internet-related problems.

## Conclusion

In summary, the prevalence of IGD and the prevalence of PIU have increased 1.6 and 1.5 times during the COVID-19 pandemic, respectively. Young people and those infected with COVID-19 are at greater risk for IGD. The pandemic has seriously changed people’s lives, and some of the changes induce problematic internet-related behaviors. It is essential to mitigate such impacts of the COVID-19 pandemic to promote healthier societies.

## Data Availability

Statistical data that support findings of this study are available in Supplementary Data. Owing to company cohort data sharing restrictions, individual-level data cannot be publicly posted. Data are, however, available from the authors upon reasonable request and with permission of KDDI Corporation.

## Acknowledgments

This research was supported by a KDDI collaborative research contract. It was also supported by Innovative Science and Technology Initiative for Security Grant Number JPJ004596, ATLA, Japan. We thank Rumi Yorizawa, Misa Murakami, Chika Hosomi, and Miho Nagata for data collection and organization. We also thank Aurelio Cortese for helpful discussion.

